# Pharmacokinetics of dexamethasone in tuberculosis meningitis

**DOI:** 10.1101/2025.07.14.25331510

**Authors:** Jose M Calderin, Juan Eduardo Resendiz-Galvan, Noha Abdelgawad, Angharad Davis, Cari Stek, Lubbe Wiesner, Graeme Meintjes, Robert J. Wilkinson, Paolo Denti, Sean Wasserman

**Affiliations:** Division of Clinical Pharmacology, Department of Medicine, University of Cape Town, Observatory, Cape Town, South Africa; Wellcome Discovery Research Platforms in Infection, Centre for Infectious Diseases Research in Africa, Institute of Infectious Disease and Molecular Medicine, University of Cape Town, Observatory, Cape Town, South Africa; The Francis Crick Institute, London, United Kingdom; Department of Medicine, University of Cape Town, Observatory, Cape Town, South Africa; Blizard Institute, Faculty of Medicine and Dentistry, Queen Mary University of London, London, United Kingdom; Department of Infectious Diseases, Imperial College London, London, United Kingdom; Institute for Infection and Immunity, City St George’s, University of London, London, United Kingdom; Division of Infectious Diseases and HIV Medicine, Department of Medicine, University of Cape Town, Observatory, Cape Town, South Africa

**Keywords:** Tuberculous Meningitis, HIV, Dexamethasone, Pharmacokinetics, Drug-Drug Interaction, Rifampicin, NONMEM, Lopinavir/Ritonavir

## Abstract

**Introduction:** Dexamethasone is recommended as adjunctive therapy for tuberculosis meningitis (TBM). Co-administration with rifampicin is expected to reduce dexamethasone exposure in TBM, an effect that may be more pronounced with the higher rifampicin doses currently being evaluated in clinical trials.

**Methods:** This pharmacokinetic study was nested in a randomised controlled trial comparing the safety of high-dose rifampicin (oral, 35 mg/kg; intravenous, 20 mg/kg) plus linezolid, with or without aspirin, vs standard-dose rifampicin (10 mg/kg) for adults with HIV-associated TBM. All participants received adjunctive oral dexamethasone every 12 hours starting at a dose of 0.4 mg/kg/day. Dexamethasone concentrations were measured on intensively sampled plasma on day 3 after study enrolment and analysed using nonlinear mixed-effects modelling.

**Results:** In total, 261 dexamethasone concentrations from 43 participants were available for model development. Eight (18%) participants were on efavirenz-based ART and five (11%) were on a lopinavir/ritonavir-based regimen. The median duration of rifampicin therapy at the time of pharmacokinetic sampling was 4 days (range: 0–7). Dexamethasone pharmacokinetics was best described by a one-compartment disposition model with first-order absorption and elimination. Typical oral clearance (CL/F) was 131 L/h, reduced to 11.5 L/h with concomitant lopinavir/ritonavir. High-dose rifampicin had no significant additional effect on dexamethasone pharmacokinetic parameters compared with the standard-dose.

**Conclusions:** In adults with HIV-associated TBM, there was high dexamethasone clearance, likely related to a drug-drug interaction with rifampicin. High-dose rifampicin had no additional effect on dexamethasone exposure.

**40-word summary of the article’s main point:** This pharmacokinetic analysis of dexamethasone in adults with HIV-associated tuberculosis meningitis found high oral clearance (131 L/h), likely due to a drug-drug interaction with rifampicin. High-dose rifampicin had no additional effect on dexamethasone exposure compared with standard dose.

## Introduction

Tuberculous meningitis (TBM) is associated with high mortality, particularly among people with HIV, and survivors are often left with chronic neurological disability [1]. Disease severity in TBM is driven by intracerebral inflammation, caused by a dysregulated immune response to *Mycobacterium tuberculosis*. Host inflammation is modulated by dexamethasone, which confers modest survival benefit when provided with antituberculosis drugs in randomised controlled trials [2–4]. Adjunctive dexamethasone is therefore recommended by international treatment guidelines for all adults with TBM.

Dexamethasone is a substrate of cytochrome P450 3A4 (CYP3A4) and P-glycoprotein (P-gp), making it susceptible to drug-drug interactions (DDI). Multiple doses of dexamethasone can also increase the transcription of both proteins [5]. Coadministration with rifampicin, the cornerstone of TBM treatment and a potent inducer of CYP3A4 and P-gp transporter activity, may reduce dexamethasone exposure [6,7], potentially affecting anti-inflammatory efficacy. This DDI may be more pronounced with higher rifampicin doses (≥20 mg/kg) currently being evaluated in TBM trials [8]. Additionally, antiretroviral drugs may be CYP3A4 inhibitors or inducers, further increasing the risk for dexamethasone DDI in HIV-associated TBM.

We aimed to characterise the pharmacokinetics of dexamethasone co-administered with standard (10 mg/kg) or high-dose rifampicin (35 mg/kg) among adults with HIV-associated TBM, specifically to evaluate whether use of high-dose rifampicin reduced dexamethasone exposure in this population.

## Methods

### Study design

This pharmacokinetic study was nested in the LASER-TBM trial (NCT03927313), which evaluated safety of intensified antituberculosis therapy among South African adults with HIV-associated TBM [9]. Participants were randomized within five days of starting TBM treatment to receive either the standard antituberculosis regimen containing rifampicin 10 mg/kg together with isoniazid, pyrazinamide, and ethambutol (R_10_HZE) or an experimental regimen containing high dose oral rifampicin 35 mg/kg (or intravenous 20 mg/kg) (R_35_HZE) and adding linezolid, with or without daily aspirin 1000 mg. Oral dexamethasone was administered to all participants every 12 hours for 4 weeks, starting at a dose of 0.4 mg/kg/day and tapering by 0.1 mg/kg per week, as recommended by national treatment guidelines. Experimental therapy was provided for 56 days, after which participants continued standard treatment.

### Pharmacokinetic sampling

Plasma samples were collected on day 3 (±2 days) after study enrolment pre-dose, at 0.5, 1, 2, 3, 6, and 8-10 hours following the morning dose, and at 12 hours post-evening dose. Immediately following collection, samples were processed on-site and stored at −80 °C. Dexamethasone concentrations were quantified using a validated liquid chromatography-tandem mass spectrometry assay (lower limit of quantification [LLOQ]: 0.938 ng/mL) performed at the Division of Clinical Pharmacology, University of Cape Town **(supplementary material S1)**.

### Pharmacokinetic modelling

Dexamethasone concentrations were described using nonlinear mixed-effects modelling in NONMEM v7.5.1 [10]. One- and two-compartment disposition models were tested with first-order absorption (with or without lag time or chain of transit compartments) and first-order elimination. Allometric scaling was applied for disposition parameters, testing body weight or fat-free mass (FFM)[11] as body size descriptors, with the exponents for clearance and volume fixed to 0.75 and 1, respectively [12]. Other covariates, including creatinine clearance (calculated using the Cockcroft-Gault formula [13]), age, treatment arm, time on rifampicin, and concomitant antiretroviral therapy (ART), were also assessed on pharmacokinetic parameters. Model development was guided by improvements in the objective function value (ΔOFV), goodness-of-fit plots, physiological plausibility and clinical relevance.

Random effects were included on the pharmacokinetic parameters if statistically significant, using a log-normal distribution [10]. Between-subject variability (BSV) was explored for disposition parameters, and between-occasion variability (BOV) was explored for absorption parameters and bioavailability, with an occasion defined as a dosing event and its subsequent observations.

Residual unexplained variability was described using an error model with both additive and proportional components, with the additive component constrained to be at least 20% of the assay’s LLOQ. Concentrations below the limit of quantification (BLQ) were handled using an adaptation of the M6 method proposed by Wijk et al [14]. Details of population pharmacokinetic modelling and handling of missing covariate data are presented in **supplementary material S2**.

The final population pharmacokinetics model was used to estimate dexamethasone 12-hour area under the curve (AUC_0-12h_) and maximum concentration (C_max_). Geometric mean ratios (GMR) for secondary pharmacokinetic parameters were computed for high-dose versus standard-dose rifampicin.

A *post hoc* power calculation showed that our study had >80% power at an alpha of 0.05 to detect a 40% reduction in dexamethasone exposure due to high-dose rifampicin.

## Results

### Study data

Dexamethasone concentrations from 43 individuals were available, consisting of 261 observations after excluding samples taken 12 hours post-evening dose which could not be used because the exact dose timing was unknown. 35/261 (13%) of the samples were below the assay’s lower limit of quantification, most of which were pre-dose observations. The median duration of rifampicin therapy at the time of pharmacokinetic sampling was 4 days (range: 0–7). All participants were HIV-positive, 8 (18%) of whom were on efavirenz-based ART and 5 (11%) on lopinavir/ritonavir-based ART, provided at double the standard-dose **(Table 1)**. Participants receiving high-dose rifampicin had lower body weight (57 kg; range 30–96) and thus received a lower total dexamethasone dose (9 mg; range 6–16) compared with those on standard-dose rifampicin (64 kg; range 42–107 and 12 mg; range 8–20, respectively) **(supplementary material Figure S3)**. Other baseline characteristics were similar between the groups **(supplementary material S4)**.

**Table 1.**
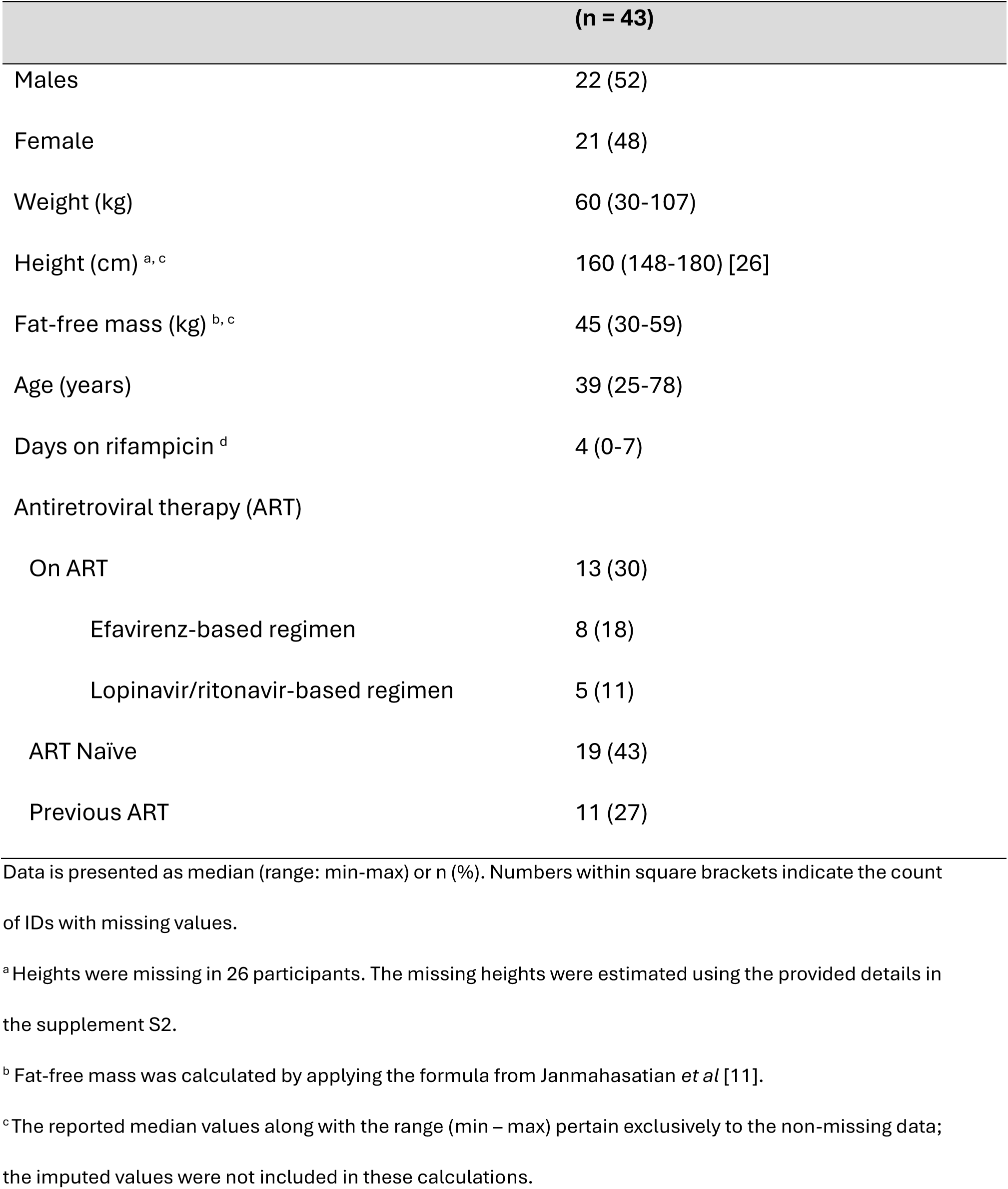
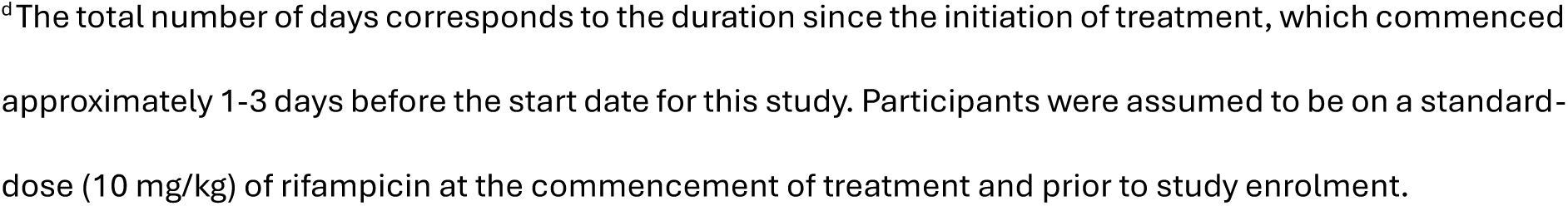
Demographic and clinical characteristics.

|  | (n = 43) |
| --- | --- |
| Males | 22 (52) |
| Female | 21 (48) |
| Weight (kg) | 60 (30-107) |
| Height (cm) <sup>a, c</sup> | 160 (148-180) [26] |
| Fat-free mass (kg) <sup>b, c</sup> | 45 (30-59) |
| Age (years) | 39 (25-78) |
| Days on rifampicin <sup>d</sup> | 4 (0-7) |
| Antiretroviral therapy (ART) |  |
| On ART | 13 (30) |
| Efavirenz-based regimen | 8 (18) |
| Lopinavir/ritonavir-based regimen | 5 (11) |
| ART Naïve | 19 (43) |
| Previous ART | 11 (27) |

Data is presented as median (range: min-max) or n (%). Numbers within square brackets indicate the count of IDs with missing values.
<sup>a</sup> Heights were missing in 26 participants. The missing heights were estimated using the provided details in the supplement S2.
<sup>b</sup> Fat-free mass was calculated by applying the formula from Janmahasatian *et al* [11].
<sup>c</sup> The reported median values along with the range (min – max) pertain exclusively to the non-missing data; the imputed values were not included in these calculations.
<sup>d</sup>The total number of days corresponds to the duration since the initiation of treatment, which commenced approximately 1-3 days before the start date for this study. Participants were assumed to be on a standard-dose (10 mg/kg) of rifampicin at the commencement of treatment and prior to study enrolment.

### Pharmacokinetic modelling

Dexamethasone pharmacokinetics was best described by a one-compartment disposition model with first-order absorption and elimination. The effect of body size was best characterized using allometric scaling based on FFM (ΔOFV = -17.7), compared with total body weight (ΔOFV = -12.3). The typical subject (FFM: 45 kg) was estimated to have a volume of distribution (V/F) of 26.6 L and an oral clearance (CL/F) of 131 L/h, which was reduced to 11.5 L/h with concomitant lopinavir/ritonavir (ΔOFV = -34.6, 1 degree of freedom, p<0.001) **(Table 2)**. High-dose rifampicin had no significant effect on dexamethasone pharmacokinetic parameters compared with the standard-dose (CL/F: ΔOFV = -1.54, p > 0.05; bioavailability: ΔOFV = -0.09, p > 0.05) **(Figure 1)**. No other covariates, including efavirenz and aspirin, significantly influenced dexamethasone pharmacokinetics.

**Figure 1.**
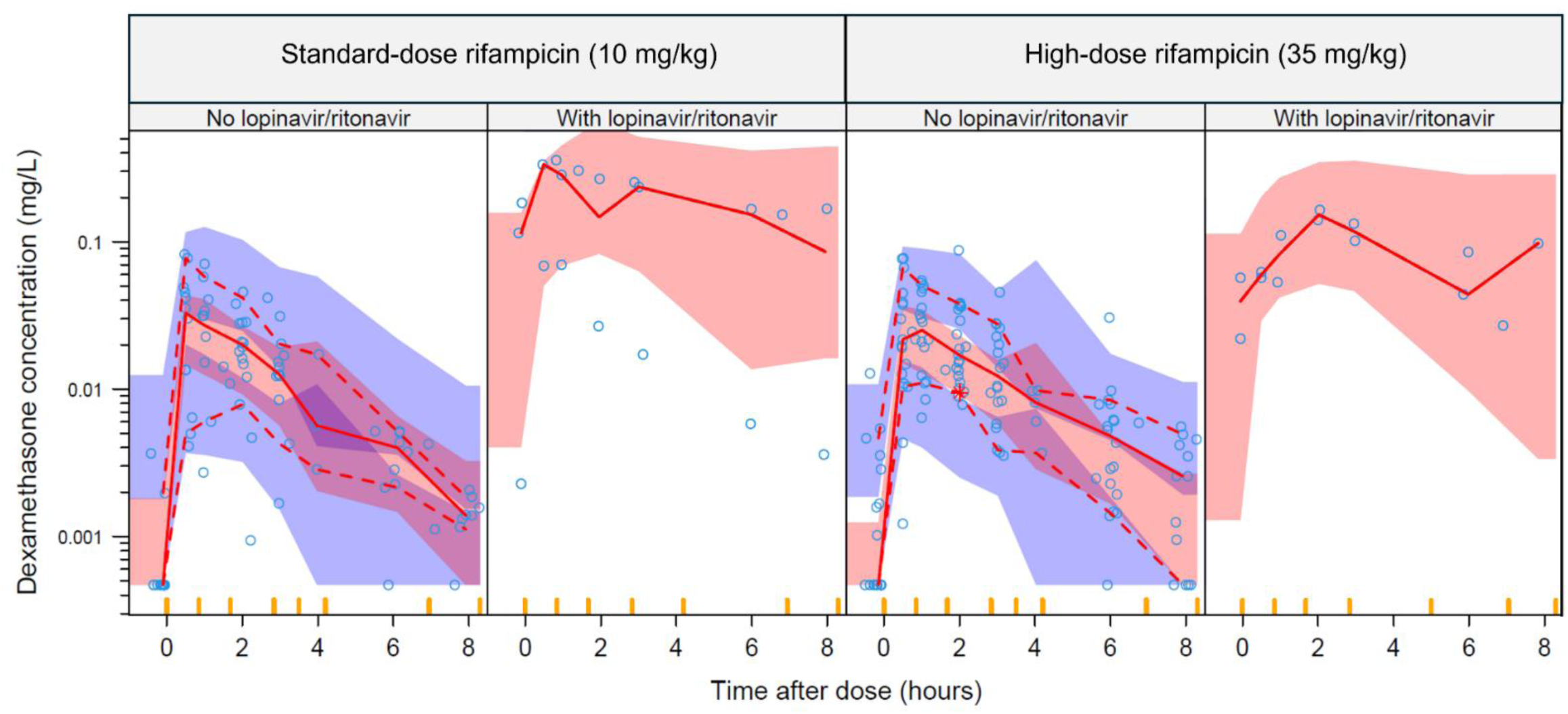
Visual predictive check of dexamethasone concentrations versus time after dose, stratified by co-administered rifampicin dose level and further stratified by lopinavir/ritonavir co-administration. Circles represent observed data. Solid and dashed lines indicate the 50th, 10th, and 90th percentiles of the observed data, while the shaded areas represent the 95% model-predicted confidence intervals for the same percentiles.

**Table 2.**
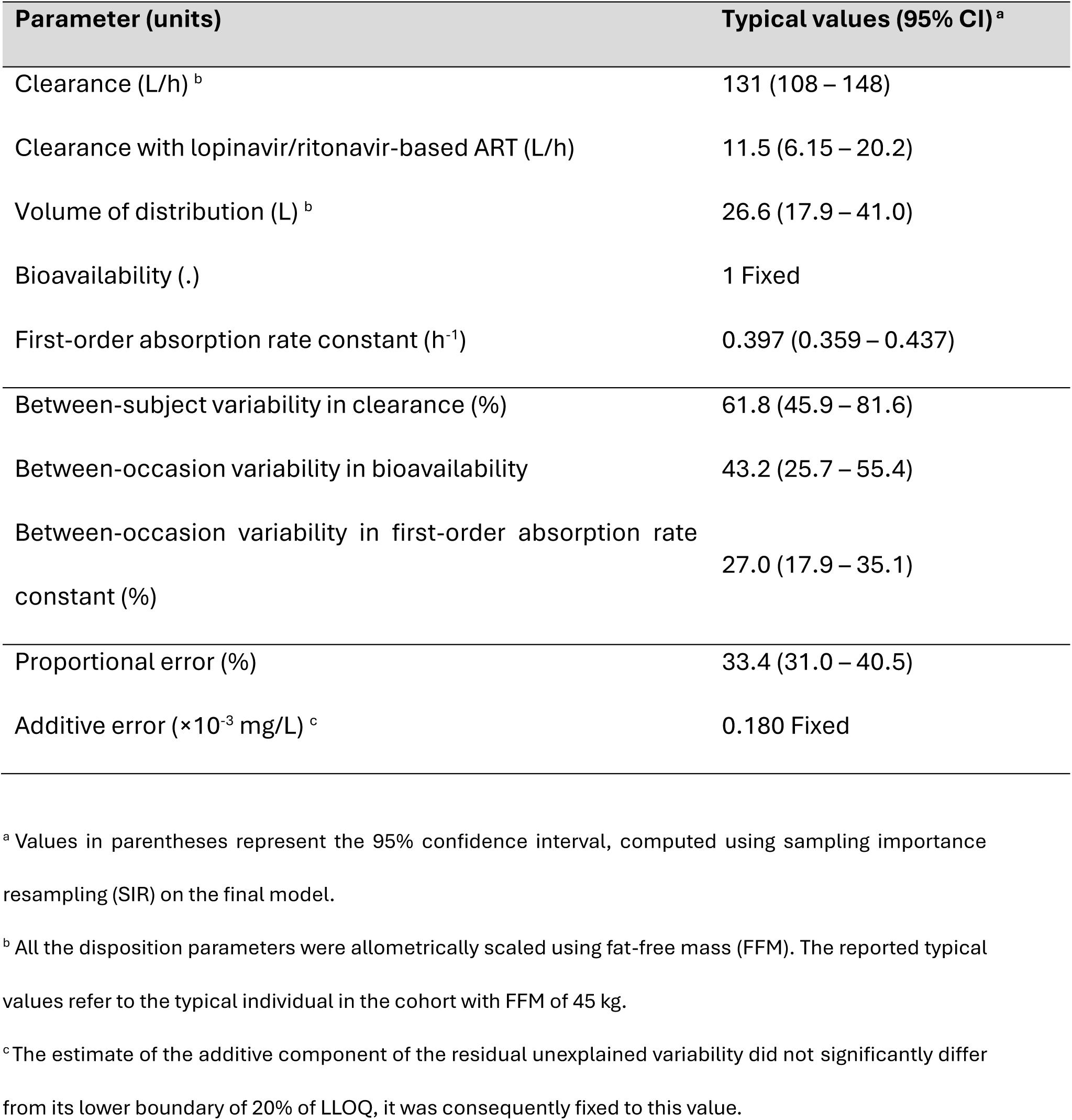
Final pharmacokinetic parameter estimates for dexamethasone.

| Parameter (units) | Typical values (95% CI) <sup>a</sup> |
| --- | --- |
| Clearance (L/h) <sup>b</sup> | 131 (108 – 148) |
| Clearance with lopinavir/ritonavir-based ART (L/h) | 11.5 (6.15 – 20.2) |
| Volume of distribution (L) <sup>b</sup> | 26.6 (17.9 – 41.0) |
| Bioavailability (.) | 1 Fixed |
| First-order absorption rate constant (h<sup>-1</sup>) | 0.397 (0.359 – 0.437) |
| Between-subject variability in clearance (%) | 61.8 (45.9 – 81.6) |
| Between-occasion variability in bioavailability | 43.2 (25.7 – 55.4) |
| Between-occasion variability in first-order absorption rate constant (%) | 27.0 (17.9 – 35.1) |
| Proportional error (%) | 33.4 (31.0 – 40.5) |
| Additive error (×10<sup>-3</sup> mg/L) <sup>c</sup> | 0.180 Fixed |

<sup>a</sup> Values in parentheses represent the 95% confidence interval, computed using sampling importance resampling (SIR) on the final model.
<sup>b</sup> All the disposition parameters were allometrically scaled using fat-free mass (FFM). The reported typical values refer to the typical individual in the cohort with FFM of 45 kg.
<sup>c</sup> The estimate of the additive component of the residual unexplained variability did not significantly differ from its lower boundary of 20% of LLOQ, it was consequently fixed to this value.

The median dexamethasone AUC_0-12h_ among participants not receiving lopinavir/ritonavir-based ART was 0.0864 mg·h/L (range: 0.0333–0.201 mg·h/L), following a median dose of 10.5 mg (range: 6–20 mg). Among those co-treated with lopinavir/ritonavir-based ART, the median AUC_0-12h_ was approximately 12 times higher, at 1.06 mg·h/L (range: 0.591–2.51 mg·h/L), after a median dose of 10 mg (range: 8–16 mg) **(Figure 2)**. Dexamethasone median AUC_0-12h_ was 0.0841 mg·h/L (range: 0.0333–1.05) in the high-dose rifampicin group and 0.111 mg·h/L (range: 0.0364–2.51) in the standard-dose group, corresponding to a GMR of 0.80 (95% CI: 0.43–1.50). Median C_max_ was 0.0275 mg/L (range: 0.0113–0.128) and 0.0347 mg/L (range: 0.00998–0.314) in the high-dose and standard-dose groups, respectively (GMR: 0.77; 95% CI: 0.47 – 1.25) **(Figure 3)**. These differences are in line with the lower median total dexamethasone dose in the high-dose rifampicin group compared to the standard-dose group (GMR: 0.83; 95% CI: 0.72– 1.00).

**Figure 2.**
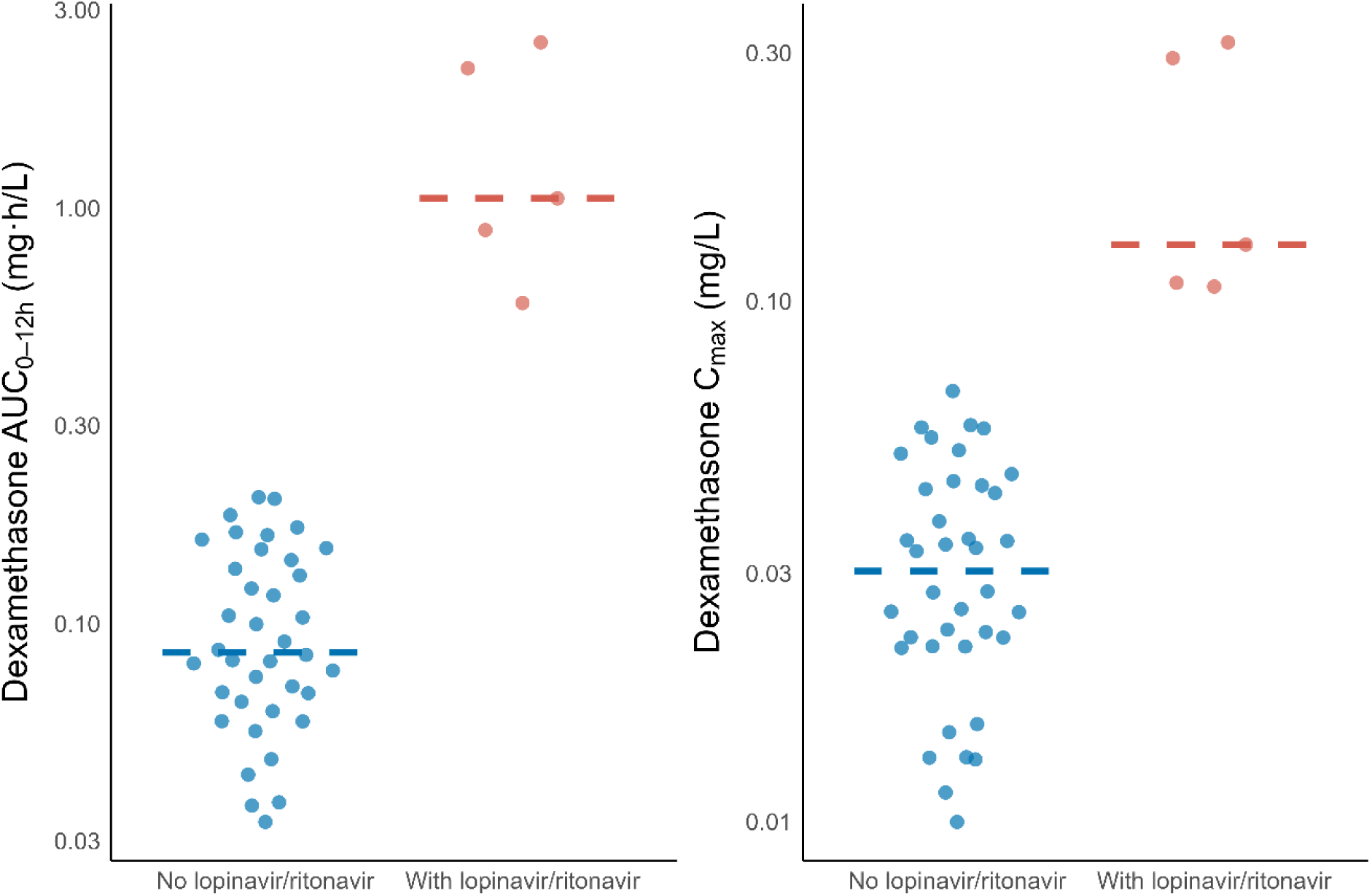
Area under the concentration–time curve from 0 to 12 hours post-dose (AUC_0-12h_) and maximum concentration (C_max_), stratified by lopinavir/ritonavir co-administration. Dots represent individual values; lines indicate the median.

**Figure 3.**
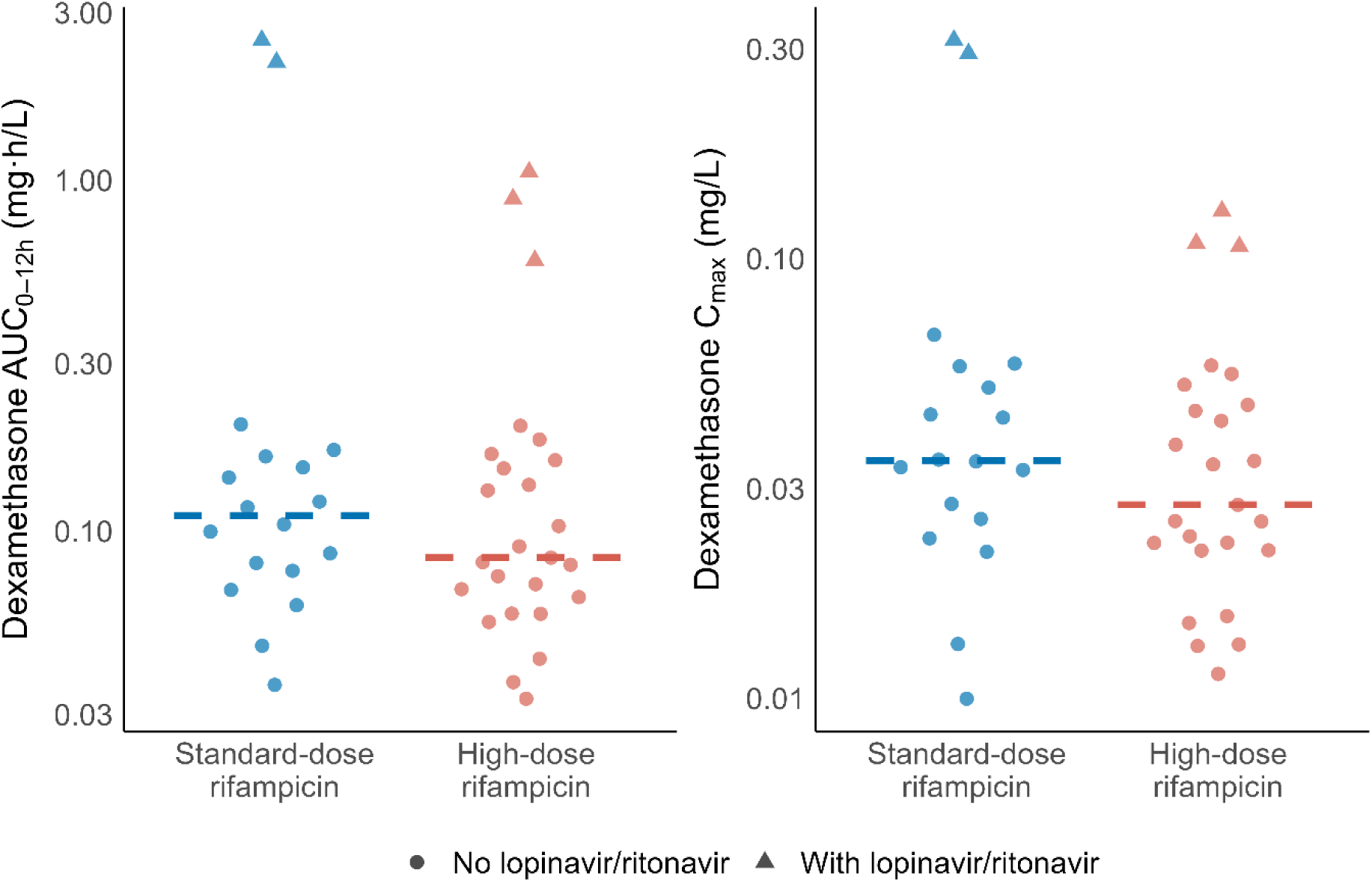
Area under the concentration–time curve from 0 to 12 hours post-dose (AUC_0-12h_) and maximum concentration (C_max_), stratified by rifampicin dose level. Dots represent individual values; lines indicate the median.

## Discussion

We characterised the pharmacokinetics of oral dexamethasone among adults with HIV-associated TBM receiving rifampicin-based treatment. In this clinical trial population, receipt of higher rifampicin doses had no additional effect on dexamethasone pharmacokinetics, and dexamethasone exposure was similar among participants receiving high-dose rifampicin (35 mg/kg) compared with standard dose rifampicin (10 mg/kg). By contrast, there was a strong inhibitory effect from lopinavir/ritonavir-based ART on dexamethasone clearance.

Limited data exist regarding dexamethasone pharmacokinetics in TBM patients, as well as its interactions with antituberculosis and antiretroviral drugs. Pharmacokinetic studies in healthy individuals receiving dexamethasone as monotherapy have reported CL/F values of 15.6 L/h following oral administration of 1.5 mg [5], and 18.1 L/h following an average intravenous dose of 5.7 mg [15]. We observed substantially higher dexamethasone CL/F of 131 L/h in our cohort of South African patients with HIV-associated TBM. This value exceeds hepatic blood flow (∼90 L/h) [16], indicating low bioavailability from significant first-pass metabolism [17].

A plausible explanation for the large dexamethasone CL/F observed in our study is a DDI with rifampicin via induction of CYP3A4 and increased expression of P-gp, enhancing dexamethasone clearance and reducing its bioavailability [18]. Previous reports support this hypothesis. A Japanese cohort (n=27) involving TB patients and healthy volunteers receiving intravenous dexamethasone reported a five-fold increase in clearance among individuals co-administered rifampicin compared with those receiving dexamethasone alone [7]. Similarly, another study involving healthy volunteers (n=16) found that, following oral administration, dexamethasone concentrations measured at 8-hours post-dose were 5- to 10-fold lower in participants co-treated with rifampicin compared with those not receiving rifampicin [6].

Indirect confirmation of the DDI with rifampicin is the much higher dexamethasone exposure observed among participants co-treated with lopinavir/ritonavir-based ART, likely due to a DDI with ritonavir, a potent CYP3A4 inhibitor [19]. Dexamethasone CL/F in this subgroup (11.5 L/h) is consistent with previous reports of dexamethasone administered alone [5], suggesting that the potent inhibitory effect of ritonavir counteracts rifampicin induction.

There is an established relationship between rifampicin concentration and CYP3A4 induction. This has been demonstrated *in vitro*, where a concentration-dependent effect from rifampicin on CYP3A4 induction was shown in liver cell lines [20], and among pulmonary TB patients (n = 24), where higher doses of rifampicin (40 mg/kg) reduced exposure of the CYP3A4 probe drug midazolam by 38% compared with standard rifampicin doses (10mg/kg) [8]. Our study had >80% power to detect such a drop in dexamethasone exposure due to high-dose rifampicin, but no trend towards faster CL/F in this group was visible in our data.

An explanation for lack of observed effect in our study is that differences in probe substrate exposure between high-dose and standard-dose rifampicin are minor compared to the large overall effect of rifampicin itself (versus no rifampicin) [21]. Illustrating this, the mean oral midazolam exposure after a single 15 mg dose without rifampicin is 170 µg·h/L [21] and, in the study of pulmonary TB patients, was reduced by 95.8% (to 7.10 µg·h/L) with 10 mg/kg rifampicin and by 97.4% (to 4.4 µg·h/L) with 40 mg/kg rifampicin [8]. This may indicate that the small additional effect of higher rifampicin doses on CYP3A4 induction does not necessarily translate into clinically significant lower dexamethasone exposures, as demonstrated in our study. The slightly lower exposures we observed in the high-dose group was attributable to receipt of lower total dexamethasone doses rather than a DDI effect from high-dose rifampicin.

Our analysis, and the probe substrate data, suggest that CYP3A4 induction from rifampicin significantly reduces dexamethasone exposure, which could potentially influence its efficacy. The recommended dexamethasone dose was established from a randomized controlled trial (n = 545) conducted in Vietnam, which demonstrated a modest survival benefit among predominantly HIV-negative patients receiving rifampicin-based TBM treatment [2]. Dexamethasone was administered intravenously in that trial, at the same dose used for oral administration in our study. Oral dexamethasone has a bioavailability of approximately 70–80% [22] resulting in lower systemic concentrations compared with intravenous dosing; this reduced bioavailability could be exacerbated because rifampicin induction is likely to have a more pronounced effect on oral dexamethasone due to extensive CYP3A4-mediated first-pass metabolism [23]. Therefore, providing oral dexamethasone alongside rifampicin to TBM patients may achieve substantially lower dexamethasone exposures than those in the Vietnam trial, with potentially reduced clinical benefit.

A more recent trial found a smaller, non-significant effect from adjunctive intravenous dexamethasone on survival among patients with HIV-associated TBM in Vietnam and Indonesia (n=520) [3]. The reason for the reduced efficacy in this population is uncertain. It may be related to secular trends in standard of care which had better outcomes compared with the original trial, reducing the relative effect from dexamethasone on survival. Another potential explanation is a DDI with efavirenz, a CYP3A4 inducer [24], resulting in increased dexamethasone clearance, as around half of participants in the dexamethasone arm (104/263) were on efavirenz-based ART [3]. In our study no differences in dexamethasone pharmacokinetics were observed among the eight participants on efavirenz-based ART, although this small sample size may have limited power to detect an effect. Additionally, since the enzyme induction pathway of efavirenz largely overlaps with that of rifampicin via activation of the pregnane X receptor [24], it is possible that no discernible effect of efavirenz was observed because participants had already been exposed to rifampicin for a median of 4 days by the time of the pharmacokinetic visit.

Our study has some limitations. First, we were unable to directly evaluate the DDI between rifampicin and dexamethasone in this population because there was no comparator group without rifampicin. However, the inhibitory effect of lopinavir/ritonavir—resulting in dexamethasone clearance values comparable to those reported historically for dexamethasone administered alone—provides indirect evidence of a DDI with rifampicin. Second, measurement of dexamethasone concentrations occurred after a median of 4 days of rifampicin exposure, prior to the full induction effect on CYP3A4. This may lead to underestimation of a DDI effect from high-dose rifampicin, which may be more pronounced after more prolonged coadministration [25]. Finally, since all patients in our study received dexamethasone orally, we were unable to estimate dexamethasone bioavailability and separate the potentially different effects of rifampicin induction on first-pass metabolism and systemic clearance.

In summary, our analysis suggests that CYP3A4 induction from rifampicin significantly reduces oral dexamethasone exposure, which could potentially influence its efficacy. Although the clinical implications are unclear—given that the target exposure for dexamethasone in TBM is unknown—prior knowledge of CYP3A4 substrate metabolism suggests that the reduction in dexamethasone exposure with oral dosing is much more pronounced than with intravenous dosing. The similar dexamethasone exposures observed between standard- and high-dose rifampicin in our study reduce the likelihood of potential harm from high-dose rifampicin in TBM due to reduced dexamethasone exposure and, consequently, a reduced anti-inflammatory effect.

## Supporting information

Supplementary Materials: Modeling methods, sample quantification, covariate imputation, and additional figures and tables.

## FUNDING

SW was supported by the Bill & Melinda Gates Foundation (INV 052110). This work was supported by Wellcome through core funding of the Wellcome Discovery research Platform in Infection (226817/Z/22/Z). AGD was supported by a UCL Wellcome Trust PhD Programme for Clinicians Fellowship (award number 175479). GM was supported by the Wellcome Trust (214321/Z/18/Z), and the South African Research Chairs Initiative of the Department of Science and Technology and National Research Foundation (NRF) of South Africa (64787). RJW receives support from the Francis Crick Institute which is funded by Wellcome (CC2112), Cancer Research UK (CC2112) and UK Research and Innovation (CC2112). He also receives support from NIH (R01145436) and in part from the NIHR Biomedical Research Center of the Imperial College Healthcare NHS Trust. The University of Cape Town Clinical PK Laboratory is supported in part via the Adult Clinical Trial Group (ACTG), by the National Institute of Allergy and Infectious Diseases (NIAID) of the National Institutes of Health under award numbers UM1 AI068634, UM1 AI068636, and UM1 AI106701; as well as the Infant Maternal Pediatric Adolescent AIDS Clinical Trials Group (IMPAACT), funding provided by National Institute of Allergy and Infectious Diseases (U01 AI068632), The Eunice Kennedy Shriver National Institute of Child Health and Human Development, and National Institute of Mental Health grant AI068632. The content is solely the responsibility of the authors and does not necessarily represent the official views of the sponsors. The funders had no role in study design, data collection and analysis, decision to publish, or preparation of the manuscript. For the purpose of Open Access, the authors have applied for a CC-BY public copyright license to any Author Accepted Manuscript version arising from this submission.

## CONFLICT OF INTEREST

All authors declare no conflict of interest.

## ACKNOWLEDGMENTS

Computations were performed using the facilities provided by the University of Cape Town’s ICTS High Performance Computing team (https://ucthpc.uct.ac.za/ ). We would like to thank Dr. Roeland Wasmann for his valuable assistance during the modelling process.

## AUTHOR CONTRIBUTIONS

SW conceptualised the study with methodology input from PD. AD and CS collected the data, provided by SW, GM and RJW. LW did the drug concentration assays. JMC, JR, and NA did the analysis and visualisation, supervised by PD. JMC and SW wrote the original draft; all coauthors reviewed and edited the manuscript. All authors approved the final draft of the manuscript. The corresponding author had full access to all the data in the study and had final responsibility for the decision to submit for publication.

## DATA AVAILABILITY

The data and model codes supporting the findings of this study are available from the corresponding author, SW, upon reasonable request.

## Notes

### Competing Interest Statement

The authors have declared no competing interest.

### Clinical Trial

NCT03927313

### Author Declarations

Ethics committee/IRB of the University of Cape Town gave ethical approval for this work (reference number 293/2018). Ethics committee/IRB of Walter Sisulu University gave ethical approval for this work (reference number 012/2019). The parent trial (LASER-TBM) is registered on ClinicalTrials.gov (identifier NCT03927313) and was approved by the South African Health Products Regulatory Authority (reference number 20180622).

