## Supplementary Materials: Modeling methods, sample quantification, covariate imputation, and additional figures and tables. for "Pharmacokinetics of dexamethasone in tuberculosis meningitis"

SUPPLEMENTARY MATERIAL

### S1. Sample quantification

Plasma dexamethasone concentrations were determined with a validated liquid chromatography-tandem mass spectrometry assay developed at the Division of Clinical Pharmacology, University of Cape Town. Samples were processed with a liquid-liquid extraction method using an isotopically labelled dexamethasone internal standard, followed by high-performance liquid chromatography with tandem mass spectrometry detection using a SCIEX 4000 instrument. Gradient chromatography was performed on an Agilent Poroshell 120 EC C18 (4.6 mm x 50 mm, 2.7 µm) analytical column with a total run time of 5 minutes. The analyte and internal standard were monitored at mass transitions of the protonated precursor 393.3 and 398.3 to the product ions 373.3 and 378.2 for dexamethasone and dexamethasone-d5, respectively. The assay was validated over the 0.938 – 240 ng/mL concentration range. The combined accuracy (%Nom) and precision (%CV) statistics of the limit of quantification, low, medium and high-quality controls (three validation batches, n=18) were between 92.8% and 100.4%, and 1.7% and 8.4%, respectively.

### S2. Pharmacokinetic modelling

Nonlinear mixed-effects modelling was conducted using NONMEM v7.5.1 to develop a population pharmacokinetic model describing dexamethasone concentrations. The first-order conditional estimation with eta-epsilon interaction (FOCE-I) algorithm was used throughout the modelling process.

One- and two-compartment disposition models were tested with first-order absorption (with or without lag time or chain of transit compartments) and first-order elimination.

We tested the effect of body size on disposition parameters using allometric scaling based on weight or fat-free mass (FFM, estimated with the formula by Janmahasatian *et al.* [1]) and the exponents for clearance and volume were fixed to 0.75 and 1, respectively [2]. The effect of other covariates including creatinine clearance (calculated using the Cockcroft-Gault formula [3]), treatment arm, and duration of concomitant rifampicin treatment, were tested on pharmacokinetic parameters. The likelihood ratio test for the drop in objective function (OFV) was used to compare nested models, assumed to be approximately $\chi^2$ distributed with n degrees of freedom, where n is the number of additional estimated parameters. A stepwise approach was used for model development; a p-value of 0.05 (ΔOFV > 3.84, 1 df) was generally employed to determine the inclusion of an additional parameter, while a value of 0.01 (ΔOFV > 6.63, 1 df) was used for its retention. Beyond statistical significance, decisions in model development were informed by diagnostic plots, including visual predictive checks, as well as physiological plausibility and clinical relevance.

Random effects were included on the pharmacokinetic parameters if statistically significant, using a log-normal distribution [4]. Between-subject variability (BSV) was explored for disposition parameters, and between-occasion variability (BOV) was explored for absorption parameters and bioavailability, with an occasion defined as a dosing event and its subsequent observations.

The residual unexplained variability (RUV) was described using an error model with both additive and proportional components, with the additive component constrained to be at least 20% of the assay's lower limit of quantification (LLOQ). Concentrations below the limit of quantification (BLQ) were handled using an adaptation of the M6 method proposed by Wijk *et al*. [5]. BLQ concentrations were imputed as half of the LLOQ, and the additive component of the RUV was increased by 50% of the respective LLOQ for these concentrations. In cases of consecutive BLQ concentrations, only one was used: the last one during the absorption phase and/or the first one during the elimination phase. Additional BLQ concentrations were excluded from parameter estimation but retained for simulation-based diagnostics. The precision of the parameter estimates in the final model, indicated by 95% confidence intervals, was evaluated through the sampling importance resampling (SIR) method [6].

#### S2.1 Missing covariates imputation

Missing heights were imputed using a methodology previously developed by Johansson and Karlsson [7]. First, patients' characteristics (sex, weight, height) from a similar population were used to develop a multiple linear regression model for heights versus weight by sex and accounting for residual variability in heights. Subsequently, this multiple linear regression model was employed in NONMEM to estimate the missing heights. The imputation model is expressed by the equation:

$$Ht_i = \beta + \alpha . Wt_i . e^{\eta_i}$$

Where Hti represents the individual height in meters, Wti is the individual weight in kilograms, β and α denote the model mean intercept and slope, respectively, and ηi is the random effect capturing the individual deviation from the mean values. The ηi values are assumed to follow a normal distribution with a mean of zero and a variance of ω2. The specific values for β and α are 1.51 and 0.00133 for females, and 1.53 and 0.00281 for males, respectively. The variance values were 0.00215 for females and 0.00170 for males.

#### S2.2 Posteriori parametric power estimation

Stochastic simulation and estimation (SSE) method [8] was performed in PsN (n = 500) using the final model to assess whether the study was sufficiently powered to detect a 40% reduction in dexamethasone exposure due to high-dose rifampicin.

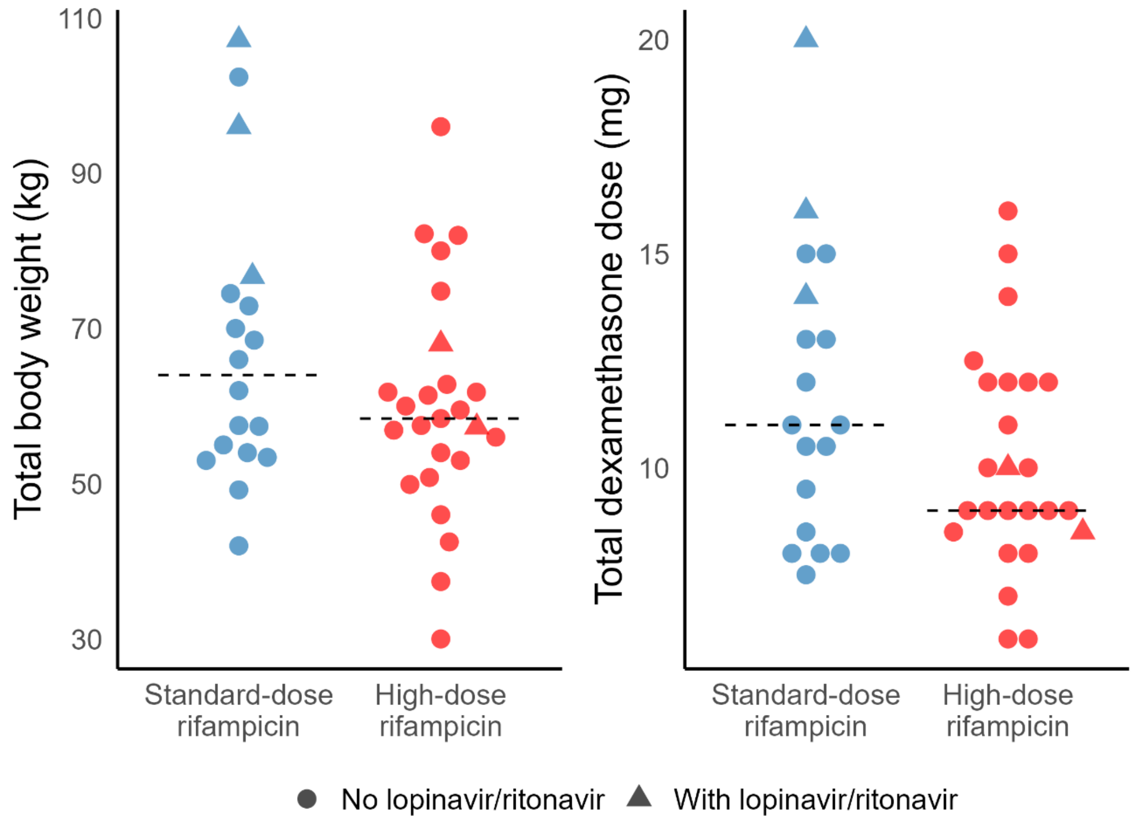


**Figure S3.** Total body weight (kg) and dexamethasone dose (mg) stratified by rifampicin dose group. Dots represent individual values; horizontal dashed lines indicate group medians.

**S4. Participant demographic and clinical characteristics by rifampicin dose group.**

| | Standard-dose rifampicin (n = 18) | High-dose rifampicin (n = 25) |
|---|---|---|
| Males | 8 (44) | 14 (56) |
| Female | 10 (56) | 11 (44) |
| Weight (kg) | 64 (42-107) | 57 (30-96) |
| Height (cm) [a, c] | 159 (149-170) [10] | 160 (148-180) [15] |
| Fat-free mass (kg) [b, c] | 46 (33-58) | 45 (30-59) |
| Age (years) | 39 (25-78) | 38 (27-56) |
| Days on rifampicin [d] | 2 (0-4) | 3 (0-7) |
| Antiretroviral therapy (ART) | | |
| On ART | 6 (33) | 7 (28) |
| Efavirenz-based regimen | 3 (17) | 5 (20) |
| Lopinavir/ritonavir-based regimen | 3 (17) | 2 (8) |
| ART Naïve | 9 (50) | 9 (36) |
| Previous ART | 3 (17) | 9 (36) |

Data is presented as median (range: Min-Max) or n (%). Numbers within brackets indicate the count of IDs with missing values.

[a] Heights were missing in 25 participants. The missing heights were estimated using the provided details in the S2.1 section, considering the sex and weight.

[b] Fat-free mass was calculated by applying the formula from Janmahasatian *et al*. [1].

[c] The reported median values along with the range (Min – Max) pertain exclusively to the non-missing data; the imputed values were not included in these calculations.

[d] The total number of days mentioned corresponds to the duration since the initiation of treatment, which commenced approximately 1-3 days before the start date for this study. It is essential to note that all participants were assumed to be on a standard-dose (10 mg/kg) of rifampicin at the commencement of treatment and prior to study enrolment.
